# The contribution of rehabilitation settings to the cost of integrated continuum-care episodes for hip fracture: a retrospective cohort study

**DOI:** 10.1101/2020.07.10.20150532

**Authors:** Davide Golinelli, Erik Boetto, Antonio Mazzotti, Simona Rosa, Paola Rucci, Elena Berti, Cristina Ugolini, Maria Pia Fantini

## Abstract

**Background:** Hip fracture (HF) requires an intensive healthcare resources utilization. Long-term morbidity related to poor fracture management is associated with a significant increase in healthcare costs. Many factors may affect the costs and outcomes in patients with HF. Using a definition of integrated Continuum-Care Episode (CCE) that encompasses the hospital phase and the post-acute rehabilitation after a surgical procedure for HF, we investigated the costs of CCEs for HF and their determinants, with particular regard to the contribution of different rehabilitation settings.

**Methods:** We conducted a retrospective observational cohort study using data extracted from administrative databases of 5094 consecutive patients hospitalized for HF in 2017, aged ≥65 years, and resident in Emilia Romagna, Italy. To evaluate the overall costs of the CCE, we calculated the acute and post-acute costs from the date of the first hospital admission to the end of the integrated CCE. The determinants of costs were investigated using generalized linear regression models.

**Results:** After adjusting for demographic and clinical characteristics, type of surgery (b=-0.340, p<0.001), and hospital bed-based rehabilitation in public or private healthcare facilities either followed by rehabilitation in a community hospital/temporary nursing home beds (b=0.372, p<0.001) or not (b=0.313, p<0.001) were the strongest determinants of costs, while rehabilitation in intermediate care facilities alone was associated with lower costs (0.163, p<0.001).

**Conclusions:** Our findings suggest that CCE cost and its variability is mainly related to the rehabilitation settings. Cost-wise, intermediate care resulted to be an appropriate setting for providing post-acute rehabilitation for HF, representing the one associated with the lower cost of the overall CCE. Therefore, intermediate care settings should be privileged when planning HF rehabilitation pathways.

## 1. INTRODUCTION

Hip fracture (HF) is a common event in the elderly that causes a growing burden on healthcare systems worldwide and requires an intensive healthcare resources utilization, especially in the first year after the fracture.^1-3^ From an epidemiological point of view, more than 300,000 hospitalizations for HF occur annually in the United States, with a 1 year mortality of almost 25%.^4^ In the United Kingdom, HF produces £ 1.1 billion of hospital costs per year, which are estimated to increase to £ 1.5 billion by 2025, when 104,000 annual cases are expected.^5^ Assuming that the age-related incidence will increase by just 1% per year, the number of HF in the world will reach approximately 8.2 million in 2050.6 HF also represents an important public health concern in Italy: in 2016, it accounted for 120,845 hospitalizations and 92,624 surgical procedures, with an annual incidence of 189.5 per 100,000 in men and of 498.4 per 100,000 in women ≥50 years old.7,8 and with relevant functional impairments in the individuals who experience it, especially the elderly.9-11

Data on HF in Italy reflect its ageing population. In Emilia Romagna, one of the largest administrative regions of North-Eastern Italy (4.4 million inhabitants), during 2011 6,368 patients aged ≥65 years were treated in regional hospitals for HF (equal to 142.8 cases per 100,000) with a clear predominance of females (75.3% of cases) and with a median age of 84 years in both sexes.12

One of the most problematic expression of population ageing is frailty,^13^ which plays an important role in reduced functional recovery and overall health status decline.^4,14,15,16^ HF in frail elderly are associated with multiple comorbidities,^17,18,19^ and several studies indicate poor functional outcomes leading to decreased mobility, less self-dependence in daily living, decreased life quality, and increased dependency and mortality rates.^20,21,22^ To reduce complications and improve functional outcomes, International guidelines recommend to perform surgery within 48 hours from hospital admission, ensure early mobilization the day after the procedure and provide a post-acute rehabilitation plan involving a multidisciplinary team in different healthcare settings.^3,23-25^

Among the possible healthcare settings addressing HF patients’ needs, in the last two decades attention was drawn to Intermediate care.^26^ Intermediate care has been developed since the late 1990s, to foster the integration between acute and primary care in several European countries.^27^ Intermediate care has both “preventive” aims (avoiding unnecessary hospitalizations and “delayed discharges”) and “rehabilitation” aims (supporting discharge and access to rehabilitation services to enforce care close to home).^28^ In fact, it is essential to ensure continuity of care by developing services and healthcare pathways that integrate hospital and primary care services.^29^ Accordingly, Intermediate care has been introduced in countries with a national and universalistic healthcare service, such as the UK and Italy, in order to provide an integrated multi-professional healthcare pathway for patients with complex/chronic needs who can be treated outside the hospital setting.^30-32^

The debate is open on what the most appropriate pathways for HF patients are. In order to define the most effective and efficient rehabilitation pathway, a Cochrane systematic review pointed to the need to evaluate all components of the process of acute care and rehabilitation together rather than separately.33 Nonetheless, there is currently no definite framework that specifies the appropriate healthcare pathway after HF.

To cope with that, Sheehan and colleagues designed a conceptual framework for an hip fracture integrated episode of care, defined as Continuum-Care Episode (CCE), that encompasses the hospital admission for the surgical procedure for HF and the following bed-based rehabilitation phase performed in different organizational settings.^34,35^

Kristen B. Pitzul and colleagues described HF patient characteristics and the most common post-acute pathways within a 1-year episode of care in Ontario, Canada, and found that similar HF patients were discharged to different post-acute settings, calling into question both the appropriateness of care delivered in the post-acute period and health system expenditures.^36^ Braithwaite et al. pointed out that almost half of the costs attributable to HF are mainly related to the costs of rehabilitation.^4^ Nonetheless, many clinical, social and organizational factors may affect the costs and outcomes of episodes of care in patients with HF. It seems therefore of interest to investigate whether rehabilitation setting and other organizational factors may determine the variability of costs in the CCE of patients with HF.^34^

The aim of this study is to investigate the costs of CCEs for hip fracture and to identify their determinants in Emilia-Romagna region, Italy, with particular regard to the contribution of different rehabilitation settings.

## 2. METHODS

### 2.1 Study setting

In Italy, the National Health System (NHS) is responsible for providing ‘essential levels of care’ to the population, through autonomous planning and delivery of healthcare services in the 21 Italian Regions.

Emilia-Romagna Regional Health System includes 8 Local Health Trusts (LHT), 4 University Hospitals, 1 General Hospital Trust, and 4 Research Hospitals. In the last decade, Emilia-Romagna region has improved the management of patients with HF, issuing policies and guidelines aimed at reducing the delay of surgery and designing specific modalities for postoperative rehabilitation.^12^ Each of the 8 LHTs is responsible for adopting regional policies and guidelines and defining predetermined healthcare pathways (or CCEs) to patients with HF. However, each LHTs is autonomous in defining specific CCEs, relying on the characteristics of its territory and on the offer of available services. In particular, a determinant of the variability in CCEs for HF patients is the availability of intermediate care facilities, such as nursing homes and community hospitals (CH). CHs are bed-based intermediate care services that may be defined as “local hospitals”, staffed mainly by general practitioners (GPs), nurses, and also rehabilitation professionals (such as physiatrists and physical therapists).

### 2.2 Study population

We conducted a retrospective observational cohort study using administrative databases of Emilia-Romagna Region that were linked and anonymized for the analysis, as reported in Supplemental Table 1 (Supplementary files).

The study population consisted of consecutive patients hospitalized for HF over the year 2017, aged ≥65 years and resident in Emilia Romagna. The inclusion and exclusion criteria were those adopted by the Italian National Outcomes Program (Programma Nazionale Esiti, PNE)7 in protocol n.42 (“Surgery within 2 days after HF in the elderly”). Accordingly, all patients hospitalized with a primary or secondary diagnosis of HF (ICD-9-CM codes 820.0-820.9), in Emilia Romagna from January 1, 2017 to December 31, 2017, were included in the study with the following exceptions: admissions preceded by hospitalization with a diagnosis of HF in the previous 2 years; hospitalizations of patients aged under 65 and over 100; hospitalizations of patients not resident in Italy; patients hospitalized after transfer from another facility; hospitalizations of polytraumatized patients (Diagnosis Related Groups -DRG 484-487); patients who died within 2 days without any surgical intervention (difference between the date of death and the date of entry to the hospital equal to 0-1 days); admissions with a primary or secondary diagnosis of malignant tumor (codes ICD-9-CM 140.0-208.9) in the index hospitalization or in the 2 previous years.

### 2.3 Continuum-care episode

According to Sheenan et al. definition^34,35^, the CCE represents an integrated episode of care consisting of an acute hospital admission phase for HF (early surgery, multidimensional evaluation and early mobilization) and a post-acute phase (rehabilitation in appropriate settings and follow-up), ending with a specific event (e.g. death of patient, long-term hospitalization in elderly care facility, or hospital discharge at home) or the end date (6 months) of the time window (Figure 1).^34,35^ Therefore, the CCE includes the acute hospitalization phase for the surgical intervention for HF (coded using the Diagnosis Related Groups - DRGs - related to HF) and continues with three different types of post-acute rehabilitation: 1) *Type 1*, Hospital bed-based rehabilitation in public or private healthcare facilities (including the same hospital); 2) *Type 2*, Hospital bed-based rehabilitation in public or private healthcare facilities (including the same hospital), followed by rehabilitation in intermediate care facilities (community hospital, CH, or temporary nursing home beds); 3) *Type 3*, Rehabilitation in intermediate care facilities (CH, or temporary nursing home beds). The CCE (Figure 1) termination rules are met when the patient: a) is deceased; b) is hospitalized in a long-term elderly care facility; c) has no other hospitalizations within 2 days from the last discharge date. The CCE (Figure 1) summarizes the healthcare and rehabilitation pathways that patients with HF can face. The CCE represents the conceptual framework that allows to describe the variability in terms of integrated services provisions and costs in Emilia Romagna. Each patient was followed up for 6 months after the first hospital admission for HF.

**Figure 1.**
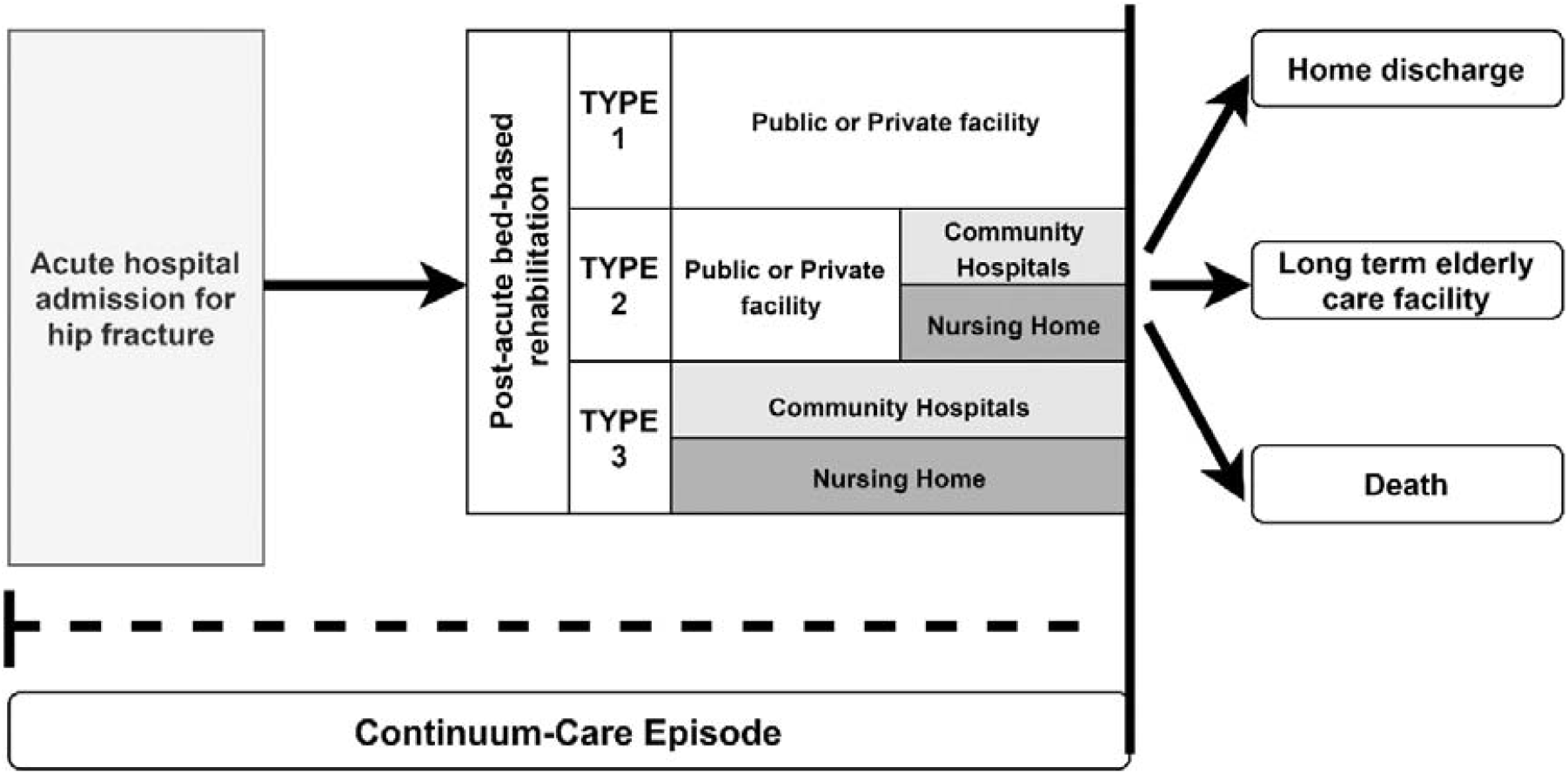
Continuum-care episode. Notes: Types of post-acute rehabilitation: 1) Type 1, Hospital bed-based rehabilitation in public or private healthcare facilities (including the same hospital); 2) Type 2, Hospital bed-based rehabilitation in public or private healthcare facilities (including the same hospital), followed by rehabilitation in intermediate care facilities (community hospital, CH, or temporary nursing home beds); 3) Type 3, rehabilitation in intermediate care facilities (CH, or temporary nursing home beds).

### 2.4 Cost analysis

The study population for the cost analysis included only patients undergoing surgery. To evaluate the overall costs of the CCE, we calculated the direct medical costs incurred by the regional healthcare system from the date of the first hospital admission to the end of the CCE, as illustrated in Figure 1. The healthcare provisions included in the CCE costs account for the acute hospitalization (i.e. DRGs) plus other costs related to: post-acute re-admissions (public or private hospital) in Orthopedics and Traumatology beds (Italian NHS bed code 036), Recovery and Rehabilitation beds (code 056), Long-term care beds (code 060), admissions to CHs, temporary admissions to nursing homes, orthopedic check-ups and outpatient rehabilitation (code R20).

### 2.5 Statistical analyses

The socio-demographic and clinical characteristics of patients and costs were summarized using descriptive statistics such as mean and standard deviation for continuous variables, and median and interquartile range for continuous asymmetric variables.

All the descriptive statistics are provided for the overall study population and by type of surgery. This was done because the type of surgical intervention is mainly influenced by the surgeons’ decision. Following the regional guidelines for surgery after HF,^12^ the decision is based on the anatomic-pathological characteristics of the fracture and on patient age and clinical conditions. This defines 4 different sub-categories of population, namely: 1) no intervention, 2) total hip replacement (THR), 3) hemiarthroplasty or partial hip replacement (PHR), 4) Open reduction and internal fixation (ORIF).

The determinants of costs were investigated using generalized linear regression models (GLM) with a gamma probability distribution and log link, to take into account the skewness of the cost distribution. The predictors of costs were chosen according to the literature and included: gender, age, morbidities (Elixhauser index), type of surgery (ORIF vs THR/PHR), timing of the surgery (over 48 hours vs. within 48 hours), an active home care program before the hospital admission (yes/no), residing in nursing home before hospital admission (yes/no), hospital length of stay (below or above the DRG threshold days), rehabilitation type (Type 1, Type 2, Type 3, vs no rehabilitation), length of rehabilitation (from 1 to 21 days vs more than 22 days)^37^, and Local Health Trust (each LHT vs Bologna LHT). The goodness of fit of the models was assessed using the following indices: the Akaike information criterion (AIC) and Bayesian information criterion (BIC). These indices have no predefined cut-offs and can only be interpreted when comparing two different models. Lower indices denote a better model fit.

## 3. RESULTS

The study population included 5094 patients aged ≥65 years with a hospital admission for HF in 2017. Table 1 shows the characteristics of the study population, by type of surgical intervention: no intervention (n = 257), THR (n = 531), PHR (n = 1793), ORIF (n = 3323). Patients with THR were on average 10 years younger (75.4 ± 7.9 years) than those with PHR (85.9 ± 6,0), ORIF (84.4 ± 7.4) and no intervention (83.7 ± 7.9). Female patients were predominant in all types of surgery (from 71.1% in THR to 78.8% in ORIF). Patients with comorbidities were 68.4% of those with THR, 56.3% and 56.5% respectively of those with PHR or ORIF and only 43.6% of those not undergoing any surgery. Patients with 3 or more comorbidities were less frequent among those undergoing THR (6.6%), while representing a large proportion of patients without any surgical intervention (27.2%). Fifteen percent died over 6 months from HF (31.5% among those without an intervention, 1.9% THR, 17.1% PHR, 14.6% ORIF).

**Table 1.** Characteristics of patients by type of intervention.

|  | No<br>intervention |  | Total hip<br>arthroplasty |  | Partial hip<br>Arthroplasty |  | Open reduction<br>and internal<br>fixation |  | TOTAL |  |
| --- | --- | --- | --- | --- | --- | --- | --- | --- | --- | --- |
|  | n=257 |  | n= 531 |  | n=1793 |  | n=3323 |  | n=5904 |  |
| **Mean age ± SD, y** | 83.7 | ± 7.9 | 75.4 | ± 5.9 | 85.9 | ± 6.0 | 84.4 | ± 7.4 | 84.0 | ± 7.5 |
| **Sex, n (col %)** |  |  |  |  |  |  |  |  |  |  |
| **Female** | 174 | (67.7) | 390 | (73.4) | 1275 | (71.1) | 2617 | (78.8) | 4456 | (75.5) |
| **Male** | 83 | (32.3) | 141 | (26.6) | 518 | (28.9) | 706 | (21.2) | 1448 | (24.5) |
| **Elixhauser**<br>**(classes),**<br>**n (col %)** |  |  |  |  |  |  |  |  |  |  |
| **0 morbidities** | 112 | (43.6) | 363 | (68.4) | 1009 | (56.3) | 1878 | (56.5) | 3362 | (56.9) |
| **1-2 morbidities** | 75 | (29.2) | 133 | (25.0) | 499 | (27.8) | 923 | (27.8) | 1630 | (27.6) |
| **3+ morbidities** | 70 | (27.2) | 35 | (6.6) | 285 | (15.9) | 522 | (15.7) | 912 | (15.4) |
| **Total** | **257** | **(4.4)** | **531** | **(9.0)** | **1793** | **(30.4)** | **3323** | **(56.3)** | **5904** | **(100)** |

Notes: SD= Standard Deviation; col=column.

Table 2 shows the empirical costs of the CCEs in € currency by type of surgery. The costs were broken down into the two components of the CCE (acute hospitalization + bed-based rehabilitation) and are provided for the total CCE (see also Figure 1). The median cost for the acute phase of patients undergoing THR and PHR intervention was the same (€ 10,079.43), corresponding to the DRG reimbursement, whereas the median costs for bed-based rehabilitation are slightly different in patients undergoing THR and PHR, respectively € 3,234.00 and € 3,198.74. While the costs for the acute phase are lower in patients undergoing ORIF (€ 6,526.13), the bed-based rehabilitation phase has a higher median cost (€ 3,622.08) compared with patients with THR and PHR.

**Table 2.** Costs for acute hospitalization, rehabilitation and total costs of CCE by type of intervention.

| Type of intervention | Acute costs (€) |  | Rehabilitation costs (€) |  | Total costs (€) |  |
| --- | --- | --- | --- | --- | --- | --- |
|  | Median | (95.0% CI) | Median | (95.0% CI) | Median | (95.0% CI) |
| No intervention | 2278.4 | (2278.4 - 2437.9) | 3640.0 | (3212.4 - 4158.0) | 3921.0 | (2437.9 - 5555.5) |
| Total hip arthroplasty | 10079.4 | (10079.4 - 10284.3) | 3234.0 | (2964.9 - 3542.0) | 11927.4 | (11633.1 - 12335.4) |
| Partial hip arthroplasty | 10079.4 | (10079.4 - 10284.3) | 3198.7 | (2990.0 - 3277.1) | 12494.2 | (12356.8 - 12643.4) |
| Open reduction and internal fixation | 6526.1 | (6526.1 - 6526.1) | 3622.1 | (3542.0 - 3770.0) | 8686.4 | (8566.2 - 8871.2) |

Notes: CI= Confidence Interval.

Table 3 shows the results of 2 GLM models predicting the costs incurred by the regional healthcare systems for HF in year 2017. The **first model (model A)** included “non-modifiable” variables such as age, gender, multimorbidity, long-term HA or residing in nursing home before hospital admission for HF and the type of intervention. In this model, the comorbidity index was associated with increased costs, while age and gender were unrelated with costs. The ORIF was less costly than hip replacement (b=-0.325, p<0.001) and, similarly, receiving home care or residing in nursing home before the hospitalization were associated with lower costs (b =-0.075, p<0.001 and b =-0.145, p<0.001, respectively). The AIC and the BIC for this model were respectively 106537 and 106590.

**Table 3.** Results of generalized linear regression models.

| Variable | Model A<br>“Not modifiable”<br>predictors<br>AIC= 106537<br>BIC=106590 |  | Model B<br>“Not modifiable” and<br>“modifiable”<br>predictors<br>AIC= 100592<br>BIC=100738 |  |
| --- | --- | --- | --- | --- |
|  | b | Pvalue | b | Pvalue |
| Male gender | -0.006 | 0.482 | -0.003 | 0.604 |
| Age | 0.000 | 0.500 | -0.001 | <0.001 |
| Number of comorbid conditions | 0.024 | <0.001 | 0.010 | <0.001 |
| ORIF vs THA/PHA | -0.325 | <0.001 | -0.340 | <0.001 |
| Active home care before hospitalization (yes vs. no) | -0.075 | <0.001 | -0.021 | 0.069 |
| Nursing home stay before hospitalization (yes vs. no) | -0.145 | <0.001 | -0.024 | 0.023 |
| Surgery after ≥48 hours from admission (yes vs. no) |  |  | 0.002 | 0.768 |
| Length of hospital stay over DRG threshold |  |  | 0.114 | <0.001 |
| Type 1 rehabilitation vs no rehabilitation |  |  | 0.313 | <0.001 |
| Type 2 rehabilitation vs no rehabilitation |  |  | 0.372 | <0.001 |
| Type 3 rehabilitation vs no rehabilitation |  |  | 0.163 | <0.001 |
| Rehabilitation duration over 22 days |  |  | 0.257 | <0.001 |
| Rehabilitation duration between 1 and 21 days |  |  | 0.000 | 0.988 |
| LHT 8 vs LHT 5 |  |  | -0.049 | <0.001 |
| LHT 7 vs LHT 5 |  |  | -0.002 | 0.855 |
| LHT 6 vs LHT 5 |  |  | -0.112 | <0.001 |
| LHT 4 vs LHT 5 |  |  | -0.055 | <0.001 |
| LHT 3 vs LHT 5 |  |  | -0.078 | <0.001 |
| LHT 2 vs LHT 5 |  |  | 0.044 | <0.001 |
| LHT 1 vs LHT 5 |  |  | 0.052 | <0.001 |

Notes: AIC=Akaike information criterion; BIC=Bayesian information criterion; b=coefficient; LHT=local health trust; THR=total hip replacement; PHR=hemiarthroplasty or partial hip replacement (PHR); ORIF= open reduction and internal fixation; CH=community hospital. Types of post-acute rehabilitation: 1) Type 1, Hospital bed-based rehabilitation in public or private healthcare facilities (including the same hospital); 2) Type 2, Hospital bed-based rehabilitation in public or private healthcare facilities (including the same hospital), followed by rehabilitation in intermediate care facilities (community hospital, CH, or temporary nursing home beds); 3) Type 3, rehabilitation in intermediate care facilities (CH, or temporary nursing home beds).

The **second model (model B)** included, in addition to model A variables, the “modifiable” variables that depend on organizational models, i.e. timing of surgery, the rehabilitation type, the hospital length of stay, the length of rehabilitation and the LHT of residence. After adjusting for case-mix variables and the type of intervention, the surgical intervention after 48 hours from the hospitalization was unrelated with costs (b=0.002, p=0.768). Type 2 rehabilitation (hospital bed-based rehabilitation in public or private healthcare facilities, followed by bed-based rehabilitation in intermediate care facilities) was associated with higher costs (b=0.372, p<0.001). Lastly, a hospital length of stay over the DRG threshold was associated with higher costs (b=0.114, p<0.001), as well as a rehabilitation lasting 22 days or more (b=0.257, p<0.001).

The inclusion of organizational variables led to a substantive improvement in the goodness of fit of the model to the data, with an AIC index of 100592 and a BIC index of 100738.

## 4. DISCUSSION

Investigating the costs of integrated CCEs for patients with HF and identifying their determinants is highly relevant for different healthcare stakeholders, given that HF is one of the conditions with the highest impact on health.^1-5^ Despite progress in surgery, HF is characterized by high mortality and a marked reduction in the quality of life in at least half of the surviving subjects; the literature suggests that HF’s clinical outcomes are more strongly associated with the demographic and clinical characteristics of the affected subjects than with the type of trauma or surgical and rehabilitative modality.^38,39^ However, less is known about the determinants of costs related to HF episodes of care. The definition of the CCE allowed us to describe the integrated healthcare pathway of patients with HF and to estimate their cost determinants.

We found that the characteristics of HF patients undergoing different types of surgery have a wide variability. First, patients with THR were ten years younger than those undergoing PHR. Moreover, we found a high prevalence of female patients in the population with HF.^39^ This results, in line with previous literature,^40-42^ confirms that older women are at higher risk of HF due to reduced bone density and other risk factors such as familial predisposition or obesity.

Our results also confirm that the choice of the type of surgery depends not only on the fracture type but also largely on patients’ clinical characteristics. When a HF leads to hip replacement surgery, PHR is the preferred option in patients with a more complicated health profile and whose physical activities are limited, while THR is usually applied in younger and healthier patients.^43^ Notably, our results indicate that the costs of patients undergoing THR and PHR do not show a substantial difference, neither for the acute part of the CCE, where the cost corresponds to the DRG, nor for the bed-based post-acute rehabilitation. As expected, ORIF costs less than hip replacement (both PHR and THR) in the acute part and in the total costs, while in these patients (and in those who are not subjected to surgery) the rehabilitation part of the CCE is more expensive.

Our findings confirmed that much of the variability in total episode costs is mainly attributable to differences in post-acute care costs, particularly to rehabilitation.^44-46^ Moreover, these costs are in line with the range found in the literature.^47^

The analysis of the determinants of CCE’s costs shows that some patients’ demographic and clinical characteristics are associated with differential costs. Gender is unrelated to CCE costs. This is interesting, as it underlines that no gender differences are present in this specific context of care. Age is weakly associated with costs. Notably, as patients’ age increases, CCE costs are reduced. This confirms what reported in literature and guidelines.^25,44,47,48^ In fact, older patients usually undergo less complex and therefore less expensive surgery, thus reducing overall costs.^43^

Increasing spending is correlated with the co-morbidity burden of geriatric HF patients, in line with previous literature.^49-52^ Johnson et al.^53^ showed that only 4.95% of HF patients presented without associated comorbidities. Many authors demonstrated that a higher comorbidity burden is associated with higher costs of care and longer length of stay for geriatric HF. Data presented here corroborates Casaletto and Gatt’s findings that patients with more comorbidities (HF) account for higher cost of care.^54^

Current geriatric HF literature supports surgical stabilization within 48 hours as a pathway to reduce morbidity, reduce mortality and preserve function.^55-58^ It is widely reported in the literature, and underlined by the guidelines, that performing the surgery within 48 hours from the hospital admission provides better clinical outcomes.^3,47^ However, our results underscore that carrying out the intervention after 48 hours from the hospital admission does not determine an increase in the overall costs of the CCE. We can therefore argue that, while the majority of the literature identifies the early intervention as an instrument to achieve better clinical and functional outcomes, it cannot be considered associated with a reduction of the total costs of the CCE.

On the contrary, active home care or being resident in a nursing home before the hospital admission for HF is associated with cost reduction. A possible explanation is that patients with these characteristics are already being treated by healthcare and social services, thus requiring less contacts with other healthcare facilities, and are also older, which has been proved to be associated with lower cost interventions.^43^

Net of other variables, hospital bed-based rehabilitation in public or private healthcare facilities, either followed by rehabilitation in a community hospital/temporary nursing home beds or not, is the rehabilitation type associated with higher overall costs of CCE. Conversely, bed-based rehabilitation performed exclusively in intermediate care facilities (CH or nursing home) is associated with overall lower costs. The duration of the rehabilitation has also a high impact on costs, in line with existing literature.^59^

This is mainly due to the lower costs attributable to the patients’ stay in intermediate care beds, which reflects the lower intensity of care required by HF patients in the post-acute phase. Net of patients’ case-mix and surgical intervention, it appears that the intermediate care organizational model contributes to the integrated episode of care for HF with a lower economic weight. Therefore, in designing appropriate integrated care pathways for this type of patient, this organizational option should certainly be taken into consideration.

### 4.1 Limitations

This study has some limitations. Our analyses are based on HF occurring in one calendar year; it is possible that variations of costs occur over time because of changes in reimbursement policies. Moreover, it is possible that the differences in costs we found reflect unmeasured differences in patient complexity, such as frailty degree, vitamin D status, and fall dynamics. Non-medical costs were not included in the analyses because they were not available. Also, in the present study we did not focus on the clinical outcomes of HF patients, as evaluating HF functional outcomes on a large retrospective cohort such as the one present here was not within the specific objectives.

## 5. CONCLUSIONS

In this study we investigated the determinants of costs of integrated continuum-care episodes for elderly patients with hip fracture. Our findings suggest that, after adjusting for patients’ case-mix and type of surgical intervention, the type of rehabilitation results to be the main determinant of overall costs, and that bed-based rehabilitation performed in intermediate care facilities is associated with lower overall spending. Cost-wise, intermediate care organizational settings should be privileged when planning hip fracture rehabilitation pathways.

## Data Availability

N/A

## AUTHORS STATEMENTS

### ETHICAL APPROVAL

The study has been approved by the AVEC (Comitato Etico Indipendente di Area Vasta Emilia Centro) research ethics committee board, ID 233/2019/OSS/AOUBo on April 17, 2019. Data used in this research was obtained from the Regional Healthcare Information System which includes detailed information on the use of healthcare services by all regional patients, with the patient as our unit of observation. The study, based on routine administrative information, was carried out in conformity with the regulations on data management of the Emilia-Romagna Region and with Italian privacy law. Administrative data were anonymised by an ad-hoc service of the Regional Department of Health and the University of Bologna have no possibility to retrospectively identify individuals included in the study.

